# Effects of iron on cardiovascular, kidney and safety outcomes in patients with chronic kidney disease: a systematic review and meta-analysis

**DOI:** 10.1101/2024.03.28.24305010

**Authors:** Bernard Chan, Amanda Varghese, Sunil V Badve, Roberto Pecoits-Filho, Murilo Guedes, Clare Arnott, Rebecca Kozor, Emma O’Lone, Min Jun, Sradha Kotwal, Geoffrey A Block, Glenn M. Chertow, Scott D Solomon, Muthiah Vaduganathan, Brendon L. Neuen

## Abstract

**Background and aims:** Heart failure and chronic kidney disease (CKD) are closely linked, with iron deficiency being highly prevalent in both conditions. Yet, major cardiovascular and nephrology guidelines offer contrasting recommendations on the use of iron. We evaluated the effects of iron versus usual care/placebo on clinical outcomes in patients with CKD.

**Methods:** We conducted a systematic review and meta-analysis of randomised trials of intravenous or oral iron in CKD (PROSPERO CRD42023453468). We searched Medline, Embase and the Cochrane Register from database inception until February 1, 2024 to identify eligible trials. We determined results overall and stratified by dialysis- and non-dialysis-requiring CKD using random effects models, with certainty of evidence assessed using the Grading of Recommendations Assessment, Development and Evaluation (GRADE) approach. The primary composite endpoint was heart failure hospitalisation or cardiovascular death.

**Results:** We identified 45 trials that met our inclusion criteria. Compared to usual care/placebo, iron reduced the risk of the primary composite endpoint (1659 events; RR 0.84, 95% CI 0.75-0.94; moderate certainty) an effect consistent across dialysis and non-dialysis requiring CKD (P-heterogeneity=0.70). The effect on the primary endpoint appeared driven by both components of hospitalisation for heart failure (RR 0.77; 95% CI 0.61-0.96; moderate certainty) and cardiovascular death (RR 0.81; 95% CI 0.65-1.02; low certainty). The incidence of serious adverse events was lower for iron compared to usual care/placebo (RR 0.90, 95% CI 0.82-0.98; moderate certainty; P-heterogeneity=0.09).

**Conclusion:** Iron therapies may reduce the risk of heart failure or cardiovascular death in patients with CKD. Randomised trials evaluating effects of iron on clinical outcomes are needed, especially in non-dialysis CKD, with or without anaemia.

## LAY SUMMARY

Heart failure and chronic kidney disease (CKD) are closely linked, with iron deficiency being common in both conditions. Yet, the major cardiac and kidney guidelines offer differing recommendations on the use of iron. We systematically reviewed and meta-analysed the current evidence to evaluate the effects of iron in adults with chronic kidney disease. Our analysis of 45 randomised trials demonstrated that iron therapies may reduce the risk of heart failure or cardiovascular death in patients with CKD, an effect consistent across dialysis and non-dialysis CKD. Further randomised trials evaluating effects of iron on clinical outcomes are needed, especially in non-dialysis CKD, with or without anaemia.

Iron is an essential trace element with biologic roles in haemoglobin synthesis, cellular function, and oxygen transport. Approximately one-third of patients with chronic kidney disease (CKD) have relative or absolute iron deficiency(1); both of which are associated with reduced health-related quality of life, cardiovascular events, and death, independent of anaemia.(2, 3) In patients with advanced CKD, iron deficiency and reduced erythropoietin synthesis are key drivers of anaemia. Among patients with kidney failure, iron deficiency and anaemia are virtually ubiquitous, owing to the effects of blood loss and chronic inflammation; chronic inflammation leads to upregulation of hepcidin and diminished intestinal iron absorption as well as sequestration of iron in reticuloendothelial cells.(4) Therefore, in patients with dialysis- and non-dialysis-requiring CKD, iron is frequently administered to manage anaemia and reduce the need for, and/or dosing of, erythropoiesis stimulating agents (ESAs).

In patients with heart failure with reduced ejection fraction who are iron deficient, multiple randomised, placebo-controlled trials have indicated that intravenous iron administration reduces hospitalisation for heart failure and improves functional status and quality of life.(5) As a result, clinical practice guidelines from the European Society of Cardiology and other expert scientific statements recommend intravenous iron to improve functional status and clinical outcomes in patients with heart failure with reduced ejection fraction *independent of anaemia*.(6) In contrast, kidney disease-focused guideline development groups, including Kidney Disease Improving Global Outcomes (KDIGO), recommend screening for iron deficiency only when anaemia develops, and largely focus on iron therapies as a tool *to manage anaemia*, with CKD trials mostly evaluating effects or iron on haemoglobin concentrations and ESA dosing.(7)

Inconsistent recommendations from widely accepted specialty care guidelines are noteworthy since heart failure and CKD frequently coexist, and the presence of one complicates the management of the other.(8, 9) The bi-directional relation between these two conditions suggests that iron might reduce the risk of heart failure and/or other cardiovascular events in patients with CKD. In addition to trials in heart failure that included many patients with CKD, several randomised trials of iron therapies in primary CKD populations have been conducted, comparing iron versus placebo or usual care, newer versus older generation formulations, intravenous versus oral supplementation, and higher versus lower doses of oral iron. To date, these results have not been systematically evaluated or quantitatively synthesised.

We therefore conducted a systematic review and meta-analysis to evaluate the effects of iron therapies on cardiovascular, kidney and safety outcomes in patients with dialysis- and non-dialysis-requiring CKD.

## METHODS

We conducted and reported this systematic review and meta-analysis in accordance with the Preferred Reporting Items for Systematic reviews and Meta-Analyses (PRISMA) statement, with prospective registration on PROSPERO (CRD42023453468).

### Search strategy and selection criteria

We searched Medline, Embase and the Cochrane Central Register of Controlled Trials (CENTRAL) from database inception to February 1, 2024 using search terms including “iron therapies” and related phrases, the names of individual iron compounds, “chronic kidney disease,” “heart failure,” and terms related to randomised controlled trials. The full search strategy, including text words and medical subject headings, is provided in the appendix (**Table S1**).

All randomised controlled trials of adults (aged ≥18 years) with CKD comparing the effects of intravenous or oral iron, with usual care or placebo, on cardiovascular, kidney and safety outcomes were eligible for inclusion. CKD was defined as an estimated glomerular filtration rate (eGFR) <60 mL/min/1.72m^2^ or an albumin-creatinine ratio >30 mg/g. Since the majority of patients with dialysis-requiring CKD receive regular iron administration as part of usual care,(10) we included trials that evaluated different dosing strategies (e.g., proactive versus reactive iron administration) in this population. We also included trials comparing iron-based phosphate binders (i.e., sucroferric oxyhydroxide and ferric citrate coordination complex) versus usual care or placebo. Secondary aims included evaluating cardiovascular, kidney and safety outcomes reported from trials that assessed the effects of (1) newer versus older iron formulations, (2) intravenous versus oral iron, and (3) higher versus lower dose oral iron (outside of dialysis-requiring CKD), where data were available. Multi-arm trials were included by combining two or more arms receiving iron therapies (when the comparison arm was usual care or placebo), or by combining treatment arms based on the generation or dose of the iron therapy (when there was no usual care or placebo arm). Trials that focused on adults who had undergone kidney transplantation were not excluded.

### Data extraction

Two authors (BC and AV) independently screened the titles and abstracts of all identified articles and, when indicated, reviewed full-text reports to identify potentially relevant studies. Any disagreements related to the eligibility of studies were discussed and resolved with a third author (BLN). The same two authors independently extracted all data using Covidence systematic review software 2023 (Veritas Health Innovation, Melbourne, Australia) and assessed risk of bias at the study level using the Cochrane Risk of Bias 2 tool. Additionally, we contacted investigators to request additional unpublished trial data for key outcomes. Any discrepancies in data extraction were also resolved by the third author (BLN).

### Outcomes

The primary endpoint was a composite of heart failure hospitalisation and cardiovascular death. Other cardiovascular outcomes included heart failure hospitalisation, cardiovascular death, myocardial infarction (MI) and stroke. Where data were available, we also evaluated the effects of iron therapies on kidney outcomes including change in eGFR, proteinuria and kidney failure requiring dialysis. We also assessed effects on all-cause mortality and serious adverse events.

### Data analysis and synthesis

We prespecified that treatment effects on clinical outcomes were to be quantitatively synthesised using a random effects model to obtain summary treatment effect estimates expressed as relative risks with associated 95% confidence intervals (95% CI). Our preference was to use hazard ratios from time-to-first event analyses, but where these were unavailable, we used estimated treatment effects obtained from recurrent events analyses. For treatment comparisons where only the number of events and participants were reported, we calculated and pooled risk ratios to maximise the information obtained from trial-level data.

We summarised effects on eGFR and proteinuria without conducting inference tests because of substantial heterogeneity in how data on these outcomes were reported (some trials reporting values at baseline and follow-up while other trials reported change from baseline in each treatment arm), as well as imbalances in baseline values between active and control arms.

Because most patients with dialysis-requiring CKD are routinely administered iron to support erythropoiesis,(10) we conducted stratified analyses according to dialysis- and non-dialysis-requiring CKD subgroups. Between-study variation was evaluated based on P heterogeneity values obtained from a random effects model, with standard chi-squared tests for heterogeneity used to assess for differences between dialysis- and non-dialysis-requiring subgroups.

We summarised the certainty of evidence for each outcome using the GRADE (Grading of Recommendations Assessment, Development and Evaluation) approach, based on the following domains: within-study risk of bias, indirectness of evidence, unexplained heterogeneity or inconsistency of results, and imprecision of results.(11)

Statistical analyses were performed using STATA version 17.0.

## RESULTS

Our search strategy yielded 2317 records, of which 248 full texts were assessed for eligibility (**Figure S1**). We identified 45 trials that met our inclusion criteria. Overall, 18 and 26 trials enrolled participants with dialysis- and non-dialysis-requiring CKD, respectively, and one trial enrolled participants from both groups. Five trials studied participants who had heart failure with reduced ejection fraction where treatment effects in CKD subgroups were reported.(12-16) There were 26 trials that compared the effects of iron versus placebo or usual care,(12-37) eight trials of intravenous versus oral iron,(38-45) nine trials of newer versus older iron formulations,(46-54) and two trials of higher versus lower oral iron doses.(55, 56). Ten trials assessed effects on a primary clinical endpoint, while biochemical or surrogate outcomes were the primary focus in 35 trials. Details of trials comparing iron versus usual care or placebo are summarised in **Table 1**, while trials of other treatment comparisons are summarised in **Tables S1-3**.

**Table 1.**
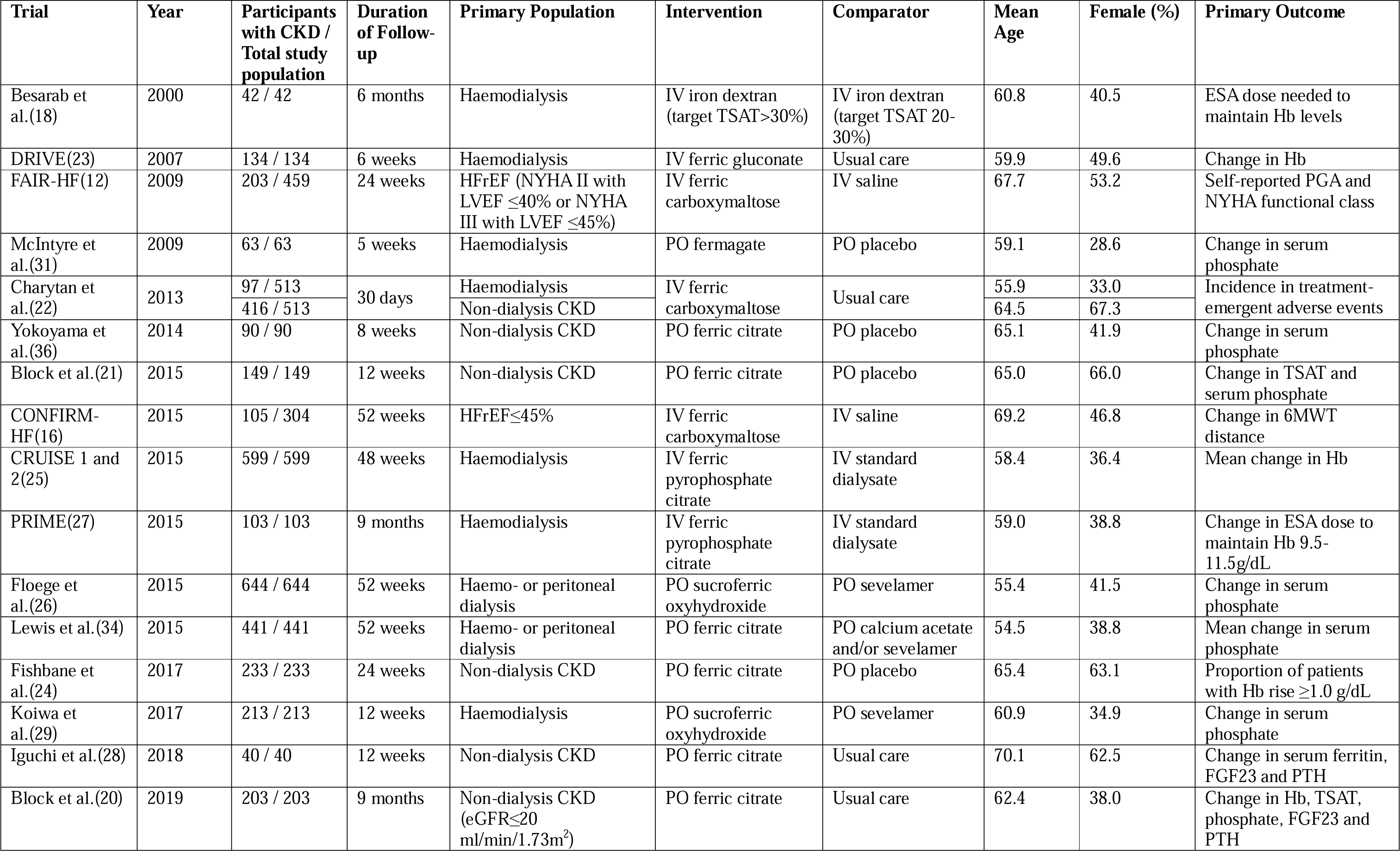
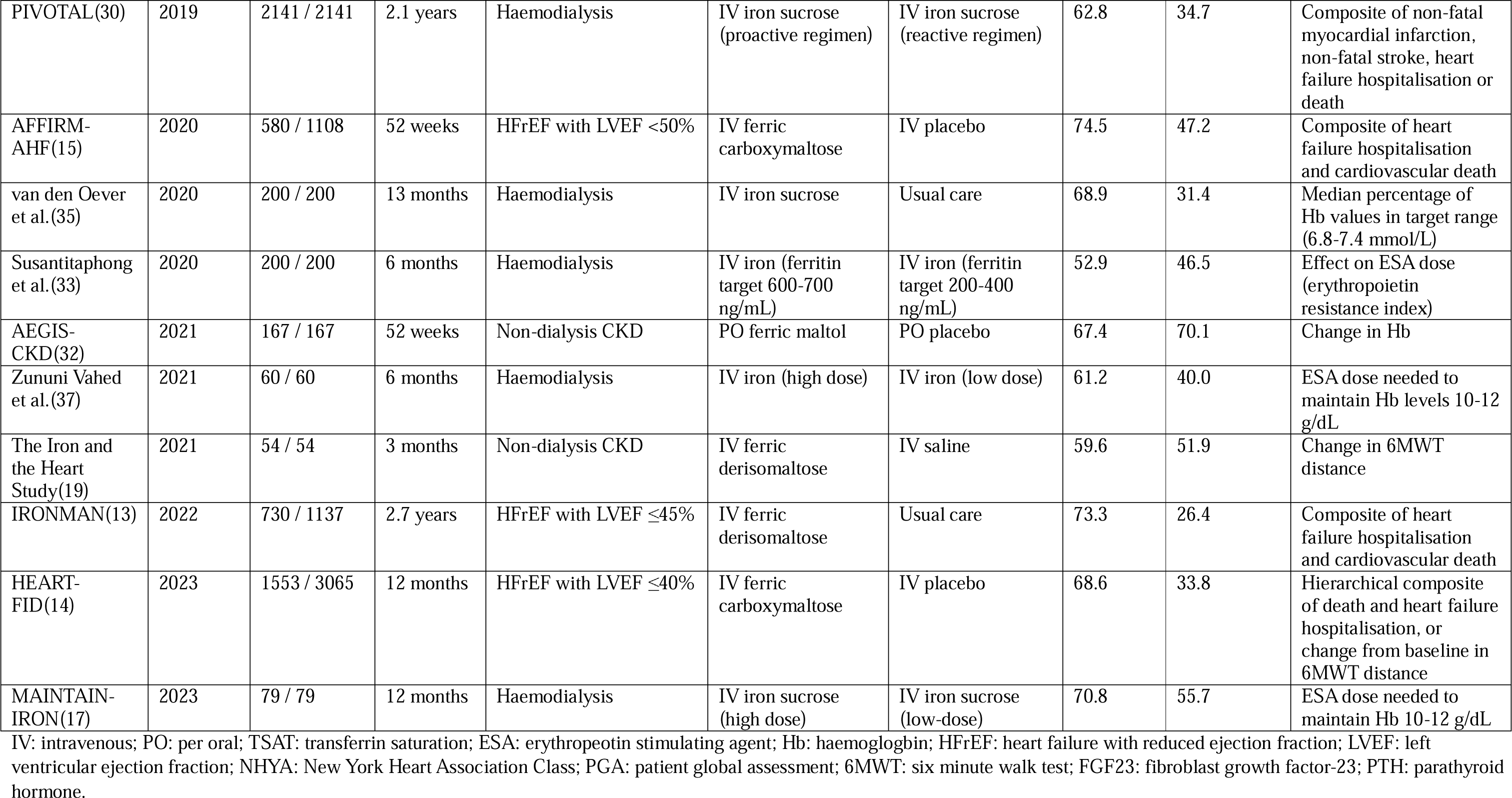
Characteristics of trials comparing iron therapies vs. usual care or placebo.

Risk of bias was low overall in the larger, event-driven trials in patients with dialysis-requiring CKD and heart failure with reduced ejection fraction (**Table S4**). However, there was substantial heterogeneity in risk of bias across other trials and for other comparisons (**Tables S4-S7**).

Overall, randomisation to iron reduced the risk of hospitalisation for heart failure or cardiovascular death by 16% (1658 events; RR 0.84, 95% CI 0.75-0.94; moderate certainty; **Figure 1**) an effect consistent across dialysis- and non-dialysis-requiring CKD subgroups (RR 0.85, 95% CI 0.75-0.96 and RR 0.81, 95% CI 0.64-1.03; P-heterogeneity=0.73). Risk reductions on the primary composite endpoint appeared driven by both components of hospitalisation for heart failure (376 events; RR 0.77; 95% CI 0.61-0.96; moderate certainty; **Figure 2**) and CV death (288 events; RR 0.81; 95% CI 0.65-1.02; low certainty; **Figure 2**), with no evidence of heterogeneity across dialysis- and non-dialysis-requiring CKD subgroups (both P-heterogeneity>0.10) For intravenous versus oral iron and newer versus older iron formulations, no clear differences were observed for these outcomes, although data were limited to a small number of trials (**Figure S2-3**).

**Figure 1.**
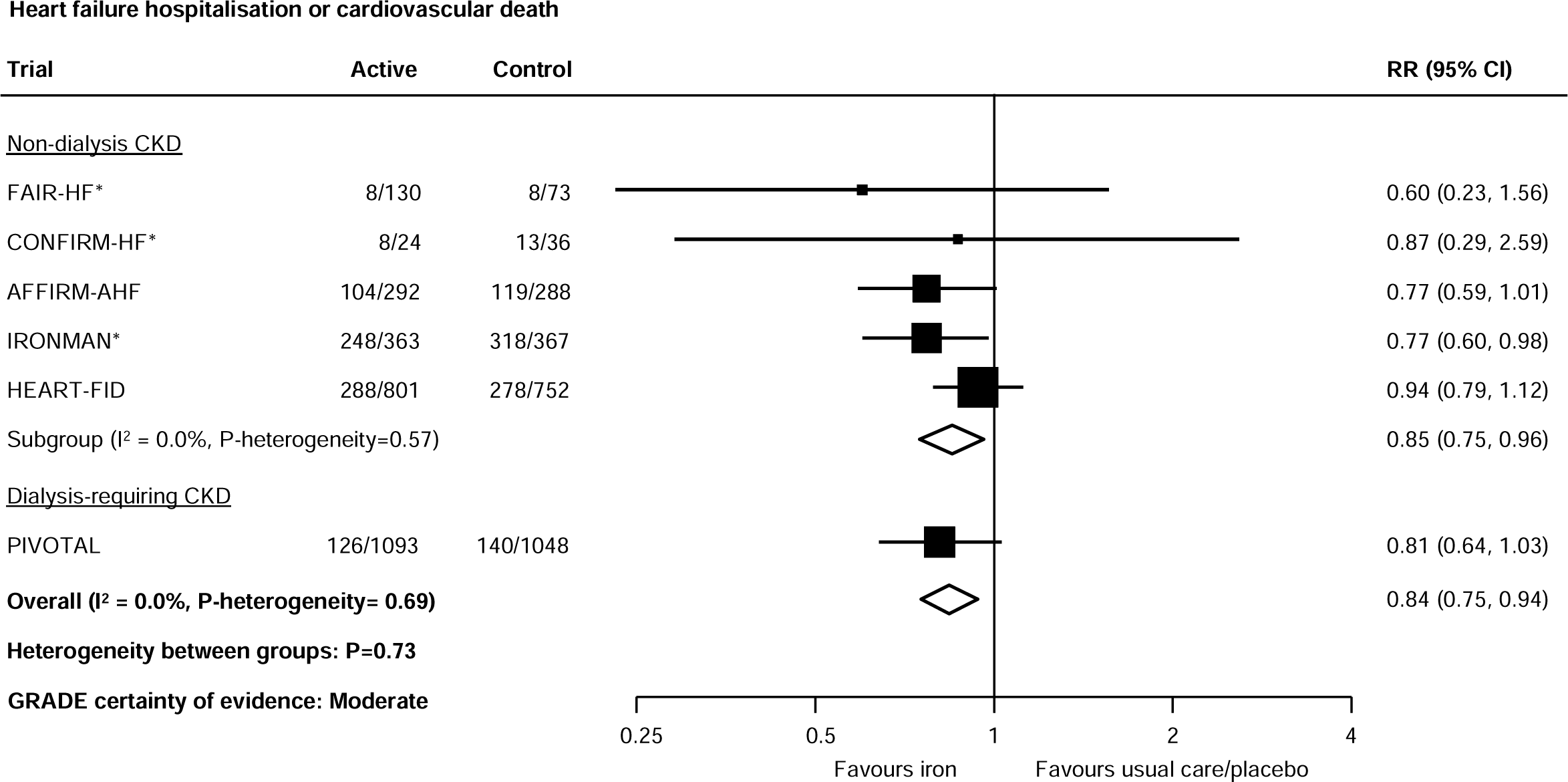
Effect of iron therapies vs. usual care or placebo on hospitalisation for heart failure or cardiovascular death in people with CKD. *First and recurrent hospitalisation for heart failure or cardiovascular death. CKD: chronic kidney disease; RR: relative risk; CI: confidence interval. GRADE: Grading of Recommendations, Assessment, Development and Evaluation.

**Figure 2.**
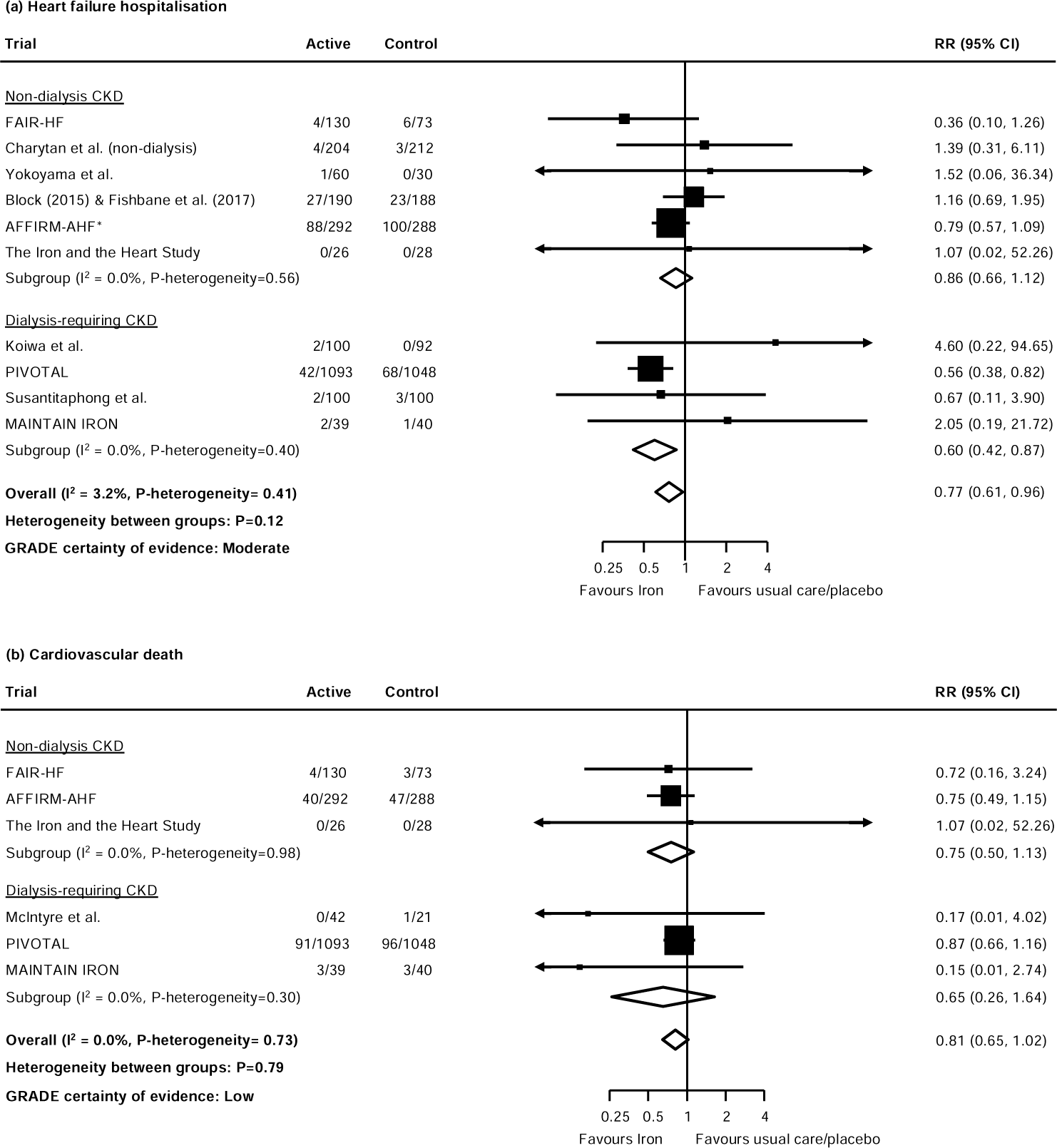
Effect of iron therapies on (A) hospitalisation for heart failure and (B) cardiovascular death in people with CKD. *First and recurrent hospitalisation for heart failure. CKD: chronic kidney disease; RR: relative risk; CI: confidence interval. GRADE: Grading of Recommendations, Assessment, Development and Evaluation.

In contrast to hospitalisation for heart failure or cardiovascular death, there were fewer myocardial infarction and stroke events recorded in the included trials. Randomisation to iron reduced the risk of MI (187 events; RR 0.66, 95% CI 0.50-0.88; low certainty; **Figure 3**), driven entirely by the PIVOTAL trial in dialysis-requiring CKD, with few events in other trials. No clear effect on stroke was observed (74 events; RR 0.91, 95% CI 0.58-1.42; very low certainty; **Figure 3**). There were too few data to reliably assess effects on MI or stroke for intravenous versus oral iron and newer versus older iron formulations (**Figures S2-3**).

**Figure 3.**
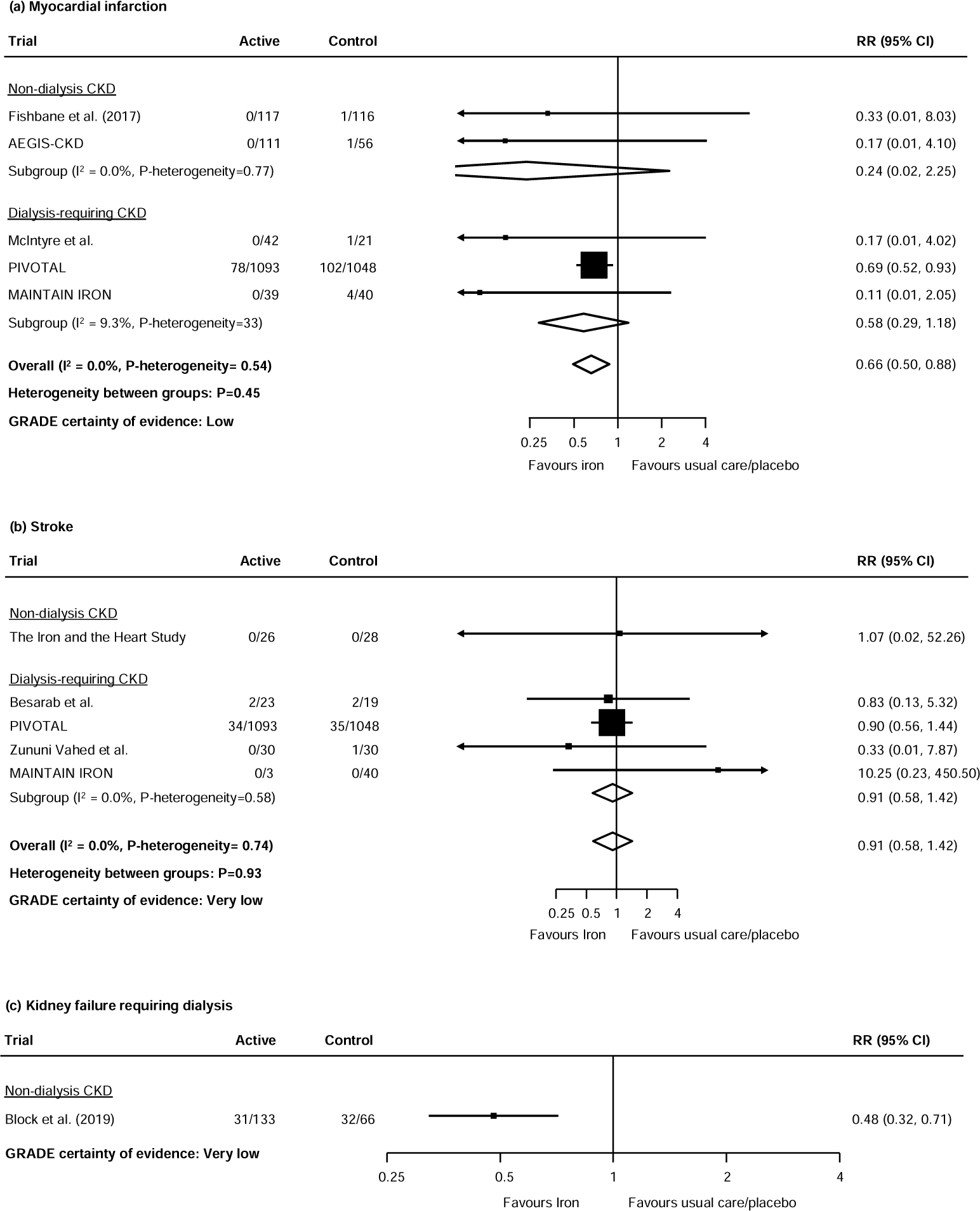
Effects of iron therapies vs. usual care or placebo on (A) myocardial infarction, (B) stroke and (C) kidney failure in people with CKD. CKD: chronic kidney disease; RR: relative risk; CI: confidence interval. GRADE: Grading of Recommendations, Assessment, Development and Evaluation.

In patients with non-dialysis-requiring CKD, randomisation to ferric citrate coordination complex reduced the risk of kidney failure based on data from one trial (63 events; RR 0.48, 95% CI 0.32-0.71; very low certainty; **Figure 3**). There were generally no effects on eGFR or proteinuria observed with iron versus usual care or placebo, but heterogeneity in data presentation and imbalances between treatment arms precluded quantitative synthesis (**Table S8-10**). For other comparisons, there were few if any data reported on kidney outcomes (**Figure S2**; **Table S11-15**).

Randomisation to iron reduced the risk of all-cause mortality (765 events; RR 0.85, 95% CI 0.74-0.98; low certainty; **Figure 4**), driven largely again by the PIVOTAL trial in dialysis-requiring CKD, although no statistical evidence of heterogeneity was observed across dialysis- and non-dialysis-requiring CKD subgroups (P-heterogeneity=0.52). Fewer serious adverse events were observed with iron compared to usual care or placebo (2,562 events, RR 0.90, 95% CI 0.82-0.98; moderate certainty; P-heterogeneity across CKD subgroups=0.09; **Figure 5**).

**Figure 4.**
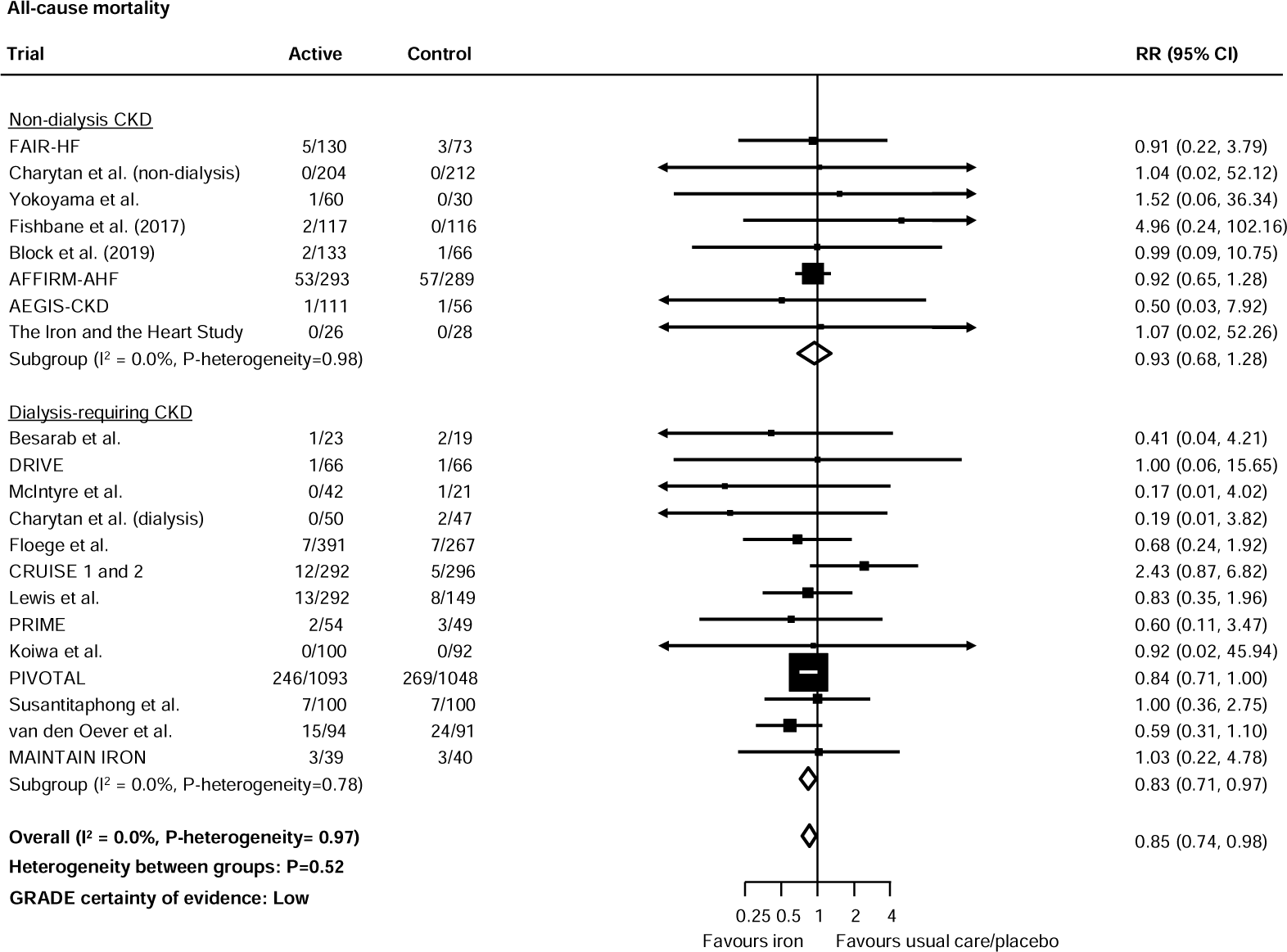
Effect of iron therapies vs. usual care or placebo on all-cause mortality in people with CKD. CKD: chronic kidney disease; RR: relative risk; CI: confidence interval. GRADE: Grading of Recommendations, Assessment, Development and Evaluation.

**Figure 5.**
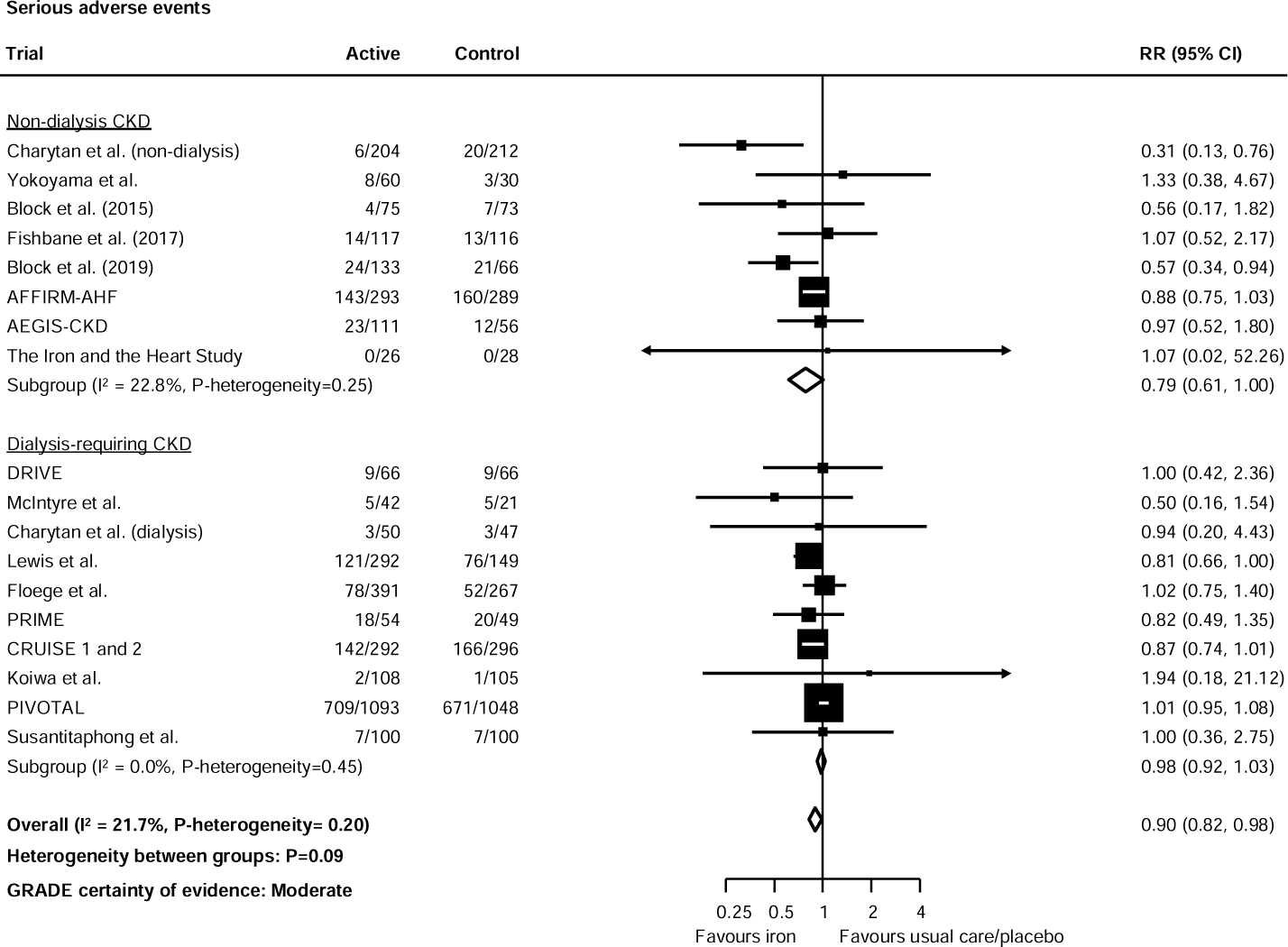
Effect of iron therapies vs. usual care or placebo on serious adverse events in people with CKD. CKD: chronic kidney disease; RR: relative risk; CI: confidence interval. GRADE: Grading of Recommendations, Assessment, Development and Evaluation.

## DISCUSSION

In this systematic review and meta-analysis, we observed that iron therapies reduced the risk of hospitalisation for heart failure or cardiovascular death compared to placebo or usual care, with consistent effects in patients with dialysis- and non-dialysis-requiring CKD. Although iron also appeared to reduce the risk of MI, kidney failure, and all-cause mortality, effects were imprecise or uncertain, often driven by a single trial not powered for these endpoints, and with very limited, if any, data in non-dialysis-requiring CKD. For comparisons of intravenous versus oral iron, newer versus older iron formulations, and higher versus lower oral iron doses, the lack of data precluded our ability to draw any reliable inferences about effects on cardiovascular events. Taken together, the data suggest that the provision of iron may yield beneficial cardiovascular effects in patients with CKD, particularly related to heart failure. Given these findings, dedicated clinical trials of intravenous or oral iron in patients with non-dialysis-requiring CKD without heart failure are warranted.

We should emphasise the fact that many of the trials included in our systematic review enrolled patients with heart failure and reduced ejection fraction; yet heart failure with preserved ejection fraction is the predominant form of heart failure in patients with CKD.(57)

In contrast to trials testing the effects of iron administration in heart failure (where we examined results in the CKD subgroup), only a single dedicated CKD trial, PIVOTAL,(58) was designed to evaluate the effects of iron on cardiovascular outcomes, despite these outcomes being consistently identified as a priority for patients and their caregivers.(59) Instead, CKD trials focused almost exclusively on correction of anaemia and changes in ESA dosing. These trials were largely motivated by concerns about ESA safety in non-dialysis-requiring CKD and economic considerations.(60, 61) The reduction in myocardial infarction, driven largely by PIVOTAL, raises the possibility that the cardiovascular benefits of iron in CKD might extend beyond reductions in heart failure and related events. But without adequately powered cardiovascular outcome trials in patients with non-dialysis CKD, effects on MI or stroke remain uncertain.

There is currently a clear distinction in recommendations between cardiology and nephrology clinical practice guidelines regarding the use of iron therapies. The European Society of Cardiology heart failure guidelines recommends intravenous iron in patients with heart failure with reduced ejection fraction who are iron deficient to improve quality of life (1A recommendation) and to reduce the risk of hospitalisation for heart failure (2A recommendation).(6) In contrast, the KDIGO 2012 guidelines for the management of anaemia in CKD recommend a trial of iron (intravenous or oral) to increase haemoglobin without starting an ESA or to reduce the dose of ESA required to maintain haemoglobin concentrations and mitigate the need for red blood cell transfusions(2C recommendation).(7) However, trials in patients with heart failure with reduced ejection fraction indicate that benefits on heart failure outcomes are *independent* of anaemia.(5)

Our findings suggest that reconsidering current recommendations on the use of iron in CKD might be appropriate and allow more patients to benefit from a therapy with a favourable benefit risk profile. A multinational study of almost 7000 patients across North America, Brazil and Europe identified significant undertreatment of iron deficiency in patients with CKD, even amongst those with anaemia.(1)

The lower incidence of serious adverse events with iron therapies versus usual care or placebo is reassuring, especially in patients with kidney failure where there has previously been concern about increased risk of infection and thrombosis.(62) Small trials, not specifically conducted in CKD, have suggested that newer iron formulations may be superior to older formulations with respect to safety;(63) however, we found no evidence that serious adverse events differed for intravenous versus oral iron and newer versus older iron formulations. Nevertheless, the overall safety profile of iron supports the feasibility of placebo-controlled outcome trials of iron therapies in CKD designed to evaluate effects on clinical outcomes.

While the available evidence suggests a potential role of iron therapies in reducing risk of worsening heart failure in CKD, this comes with the caveat that almost all the data for this outcome were derived from trials of patients with heart failure with reduced ejection fraction. Thus, it is difficult to disentangle whether these effects are driven by the presence of heart failure with reduced ejection fraction, rather than CKD per se. While there were few available data on kidney outcomes, the finding that ferric citrate coordination complex (compared to usual care) reduced the need for dialysis in a single phase 2 trial raises the possibility that iron therapies may slow the progression of kidney disease, or ameliorate signs or symptoms of uremia (or heart failure) that might prompt dialysis initiation.(64) These data, and the uncertainty they highlight, underscores the need for adequately powered randomised trials evaluating the effects of iron on functional status, health-related quality of life and clinical outcomes, especially in patients with non-dialysis-requiring CKD, with or without anaemia.

This work benefits from a pre-specified, comprehensive and systematic approach to the quantitative synthesis and interpretation of many randomised trials. We evaluated data for multiple treatment comparisons. However, some important limitations should be recognised. This was a two-stage tabular meta-analysis using mostly published data and thus we were not able to assess treatment interactions across important subgroups such as those based on transferrin saturation, serum ferritin or haemoglobin. Outside of the large event-driven outcome trials, cardiovascular events were generally not independently adjudicated, which may be particularly relevant given the clinical overlap between heart failure and CKD progression. Data for some iron products, such as ferric derisomaltose, were only available for the primary outcome. Limited data for MI, stroke and CKD progression are reflected in imprecise treatment effect estimates and limited our ability to test for heterogeneity across dialysis- and non-dialysis-requiring CKD subgroups. Accepting these limitations, our findings provide the most comprehensive assessment of the therapeutic landscape for iron therapies in CKD and underscores the need for randomised, placebo-controlled outcome trials in patients with CKD, especially those not on dialysis.

In summary, iron therapies may reduce the risk of hospitalisation for heart failure or cardiovascular death in patients with CKD. Randomised trials evaluating effects of iron on clinical outcomes are warranted, especially in non-dialysis CKD, with or without anaemia.

## Data Availability

All data produced in the present work are contained in the manuscript

## ACKNOWLEDGEMENTS

**BLN** is supported by an Australian National Health and Medical Research Council Emerging Leader Investigator Grant (grant number 2026621) and a Ramaciotti Foundation Health Investment Grant (grant number 2023HIG69). The funders had no role in the writing of the manuscript or decision to submit for publication.

