## Supplementary Appendix for "Effects of iron on cardiovascular, kidney and safety outcomes in patients with chronic kidney disease: a systematic review and meta-analysis"

Figure S1. Identification of eligible studies: PRISMA flow diagram.

Figure S2. The effects of intravenous compared to oral iron on (A) heart failure hospitalization, (B) cardiovascular death, (C) myocardial infarction, (D) stroke, (E) any cause death, and (F) serious adverse events.

Figure S3. The effects of newer vs. older generation iron products on (A) heart failure hospitalization, (B) cardiovascular death, (C) myocardial infarction, (D) stroke, (E) any cause death, and (F) serious adverse events.

Table S1. Characteristics of included trials comparing intravenous vs. oral iron therapies

Table S2. Characteristics of included trials comparing new vs. older generation iron formulations

Table S3. Characteristics of included trials comparing higher vs. lower dose oral iron therapies

Table S4. Risk of bias assessment – iron vs. usual care or placebo.

Table S5. Risk of bias assessment – intravenous vs. oral iron.

Table S6. Risk of bias assessment – newer generation iron vs. older generation iron formulations.

Table S7. Risk of bias assessment – higher vs. lower dose oral iron.

Table S8. Changes in eGFR with iron therapy vs. usual care or placebo

Table S9. Changes in eGFR with intravenous vs. oral iron

Table S10. Changes in eGFR with higher vs. lower dose oral iron

Table S11. Changes in proteinuria with iron therapy vs. usual care or placebo

Table S12. Changes in proteinuria with newer vs older generation iron

Table S13. Changes in proteinuria with higher vs. lower dose oral iron

Table S14. Changes in albuminuria with iron therapy vs. usual care or placebo

Table S15. Changes in albuminuria with newer vs older generation iron

References

PRISMA Checklist

**Figure S1. Identification of eligible studies: PRISMA flow diagram.**

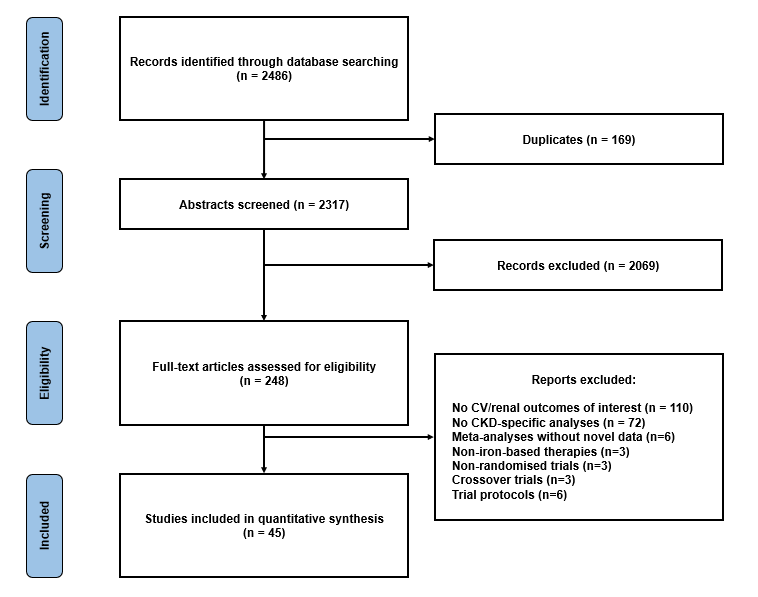

CV: cardiovascular; CKD: chronic kidney disease.

**Figure S2. The effect of intravenous compared to oral iron therapies on (A) heart failure hospitalisation, (B) cardiovascular death, (C) myocardial infarction, (D) stroke, (E) all-cause mortality, and (F) serious adverse events.**

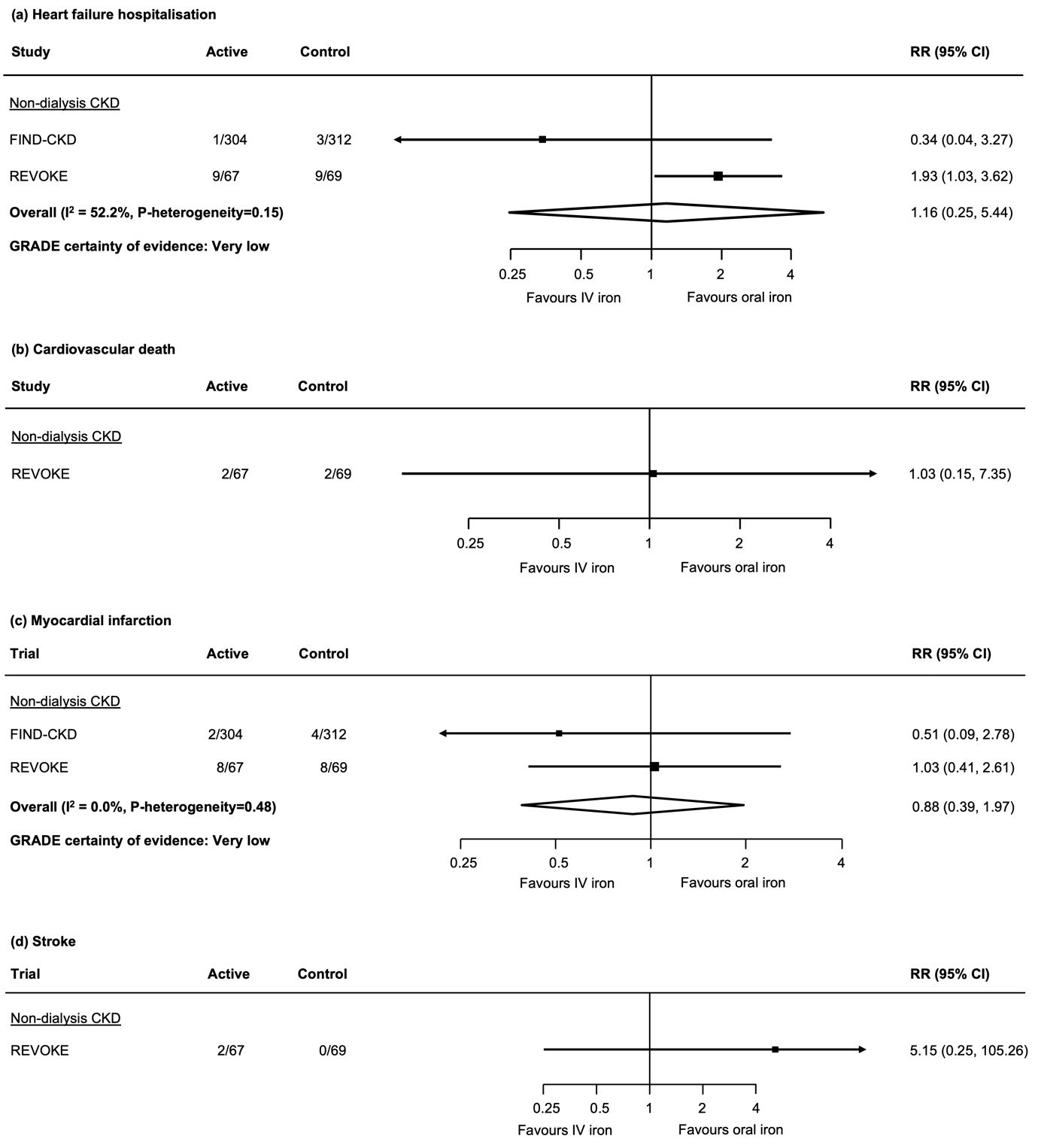

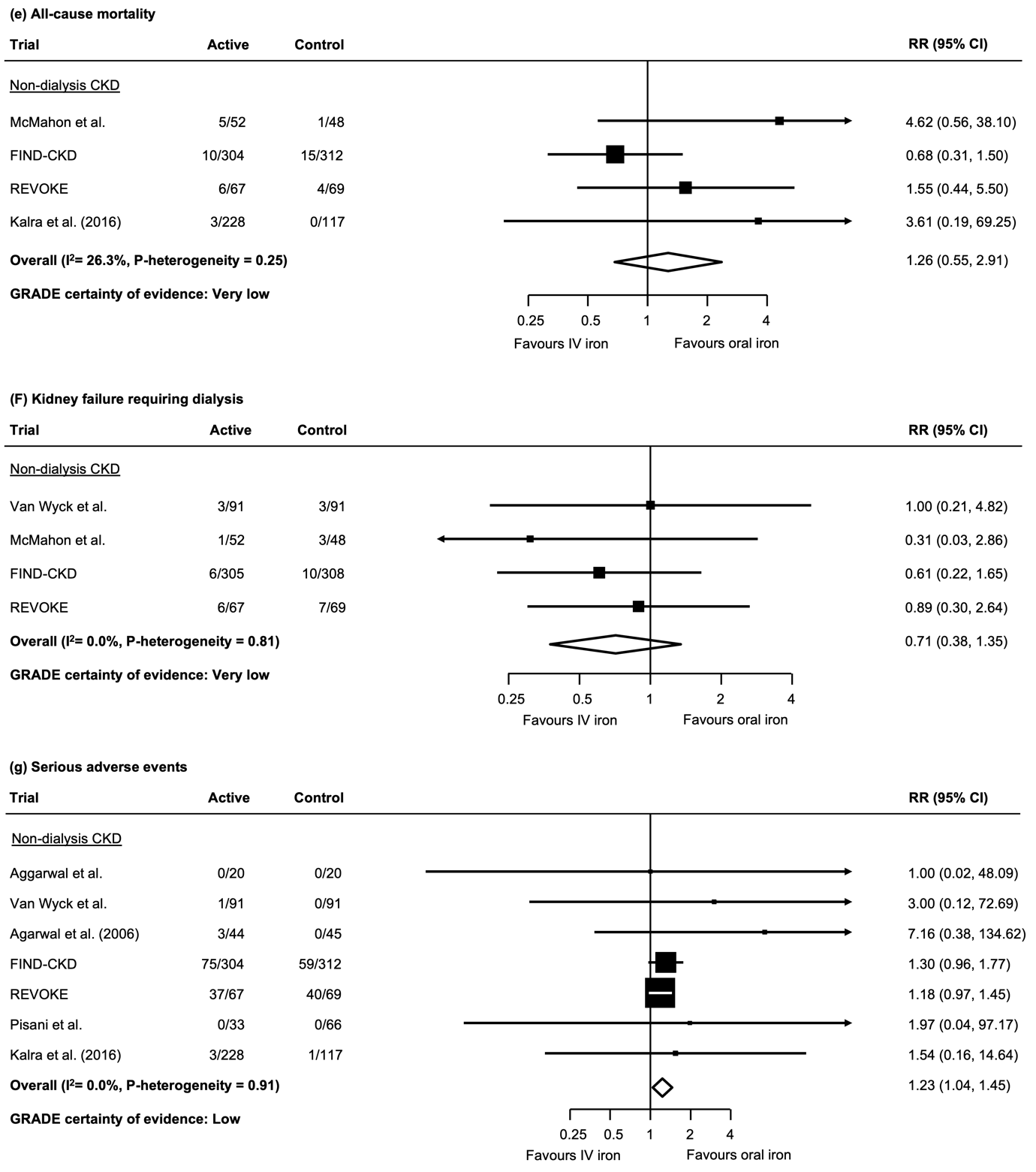

CKD: chronic kidney disease; RR: relative risk; CI: confidence interval.

**Figure S3. The effect of newer vs. older generation iron formulations on (A) heart failure hospitalization, (B) cardiovascular death, (C) myocardial infarction, (D) stroke, (E) all-cause mortality, and (F) serious adverse events.**

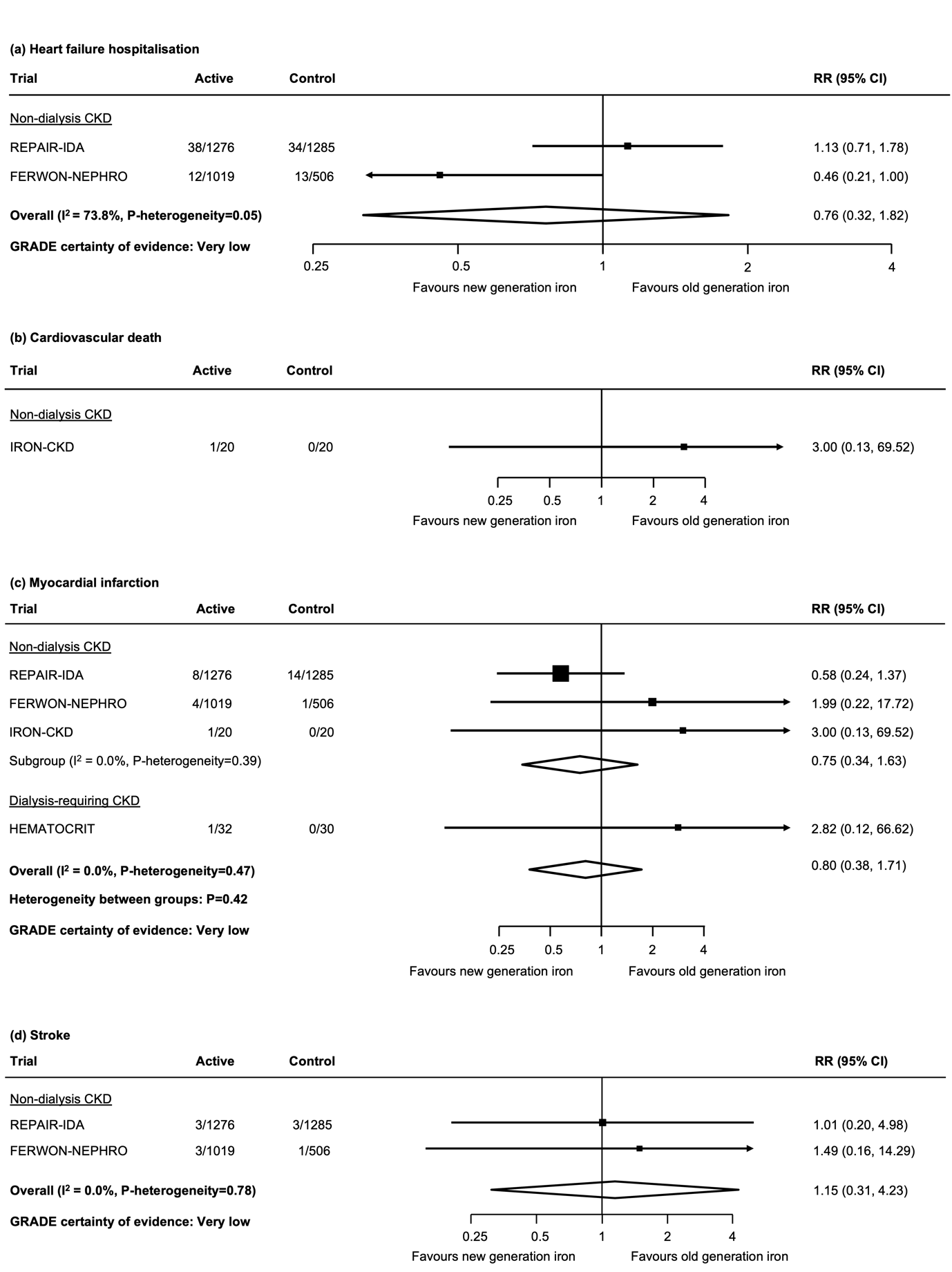

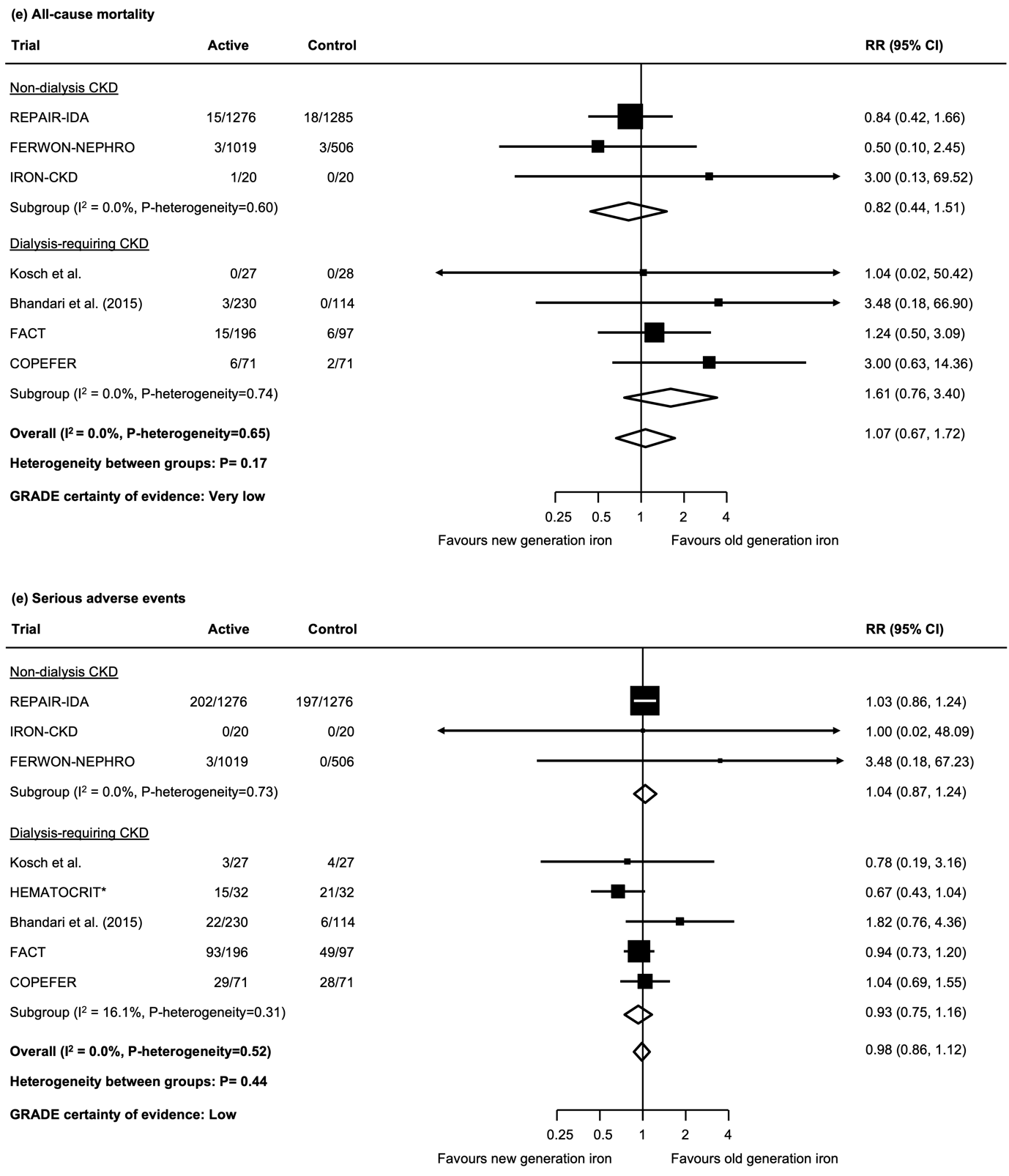

*Recurrent events analysis.

CKD: chronic kidney disease; RR: relative risk; CI: confidence interval.

**Figure S4. The effect of high vs. low dose oral iron products on (A) all-cause mortality, and (B) serious adverse events.**

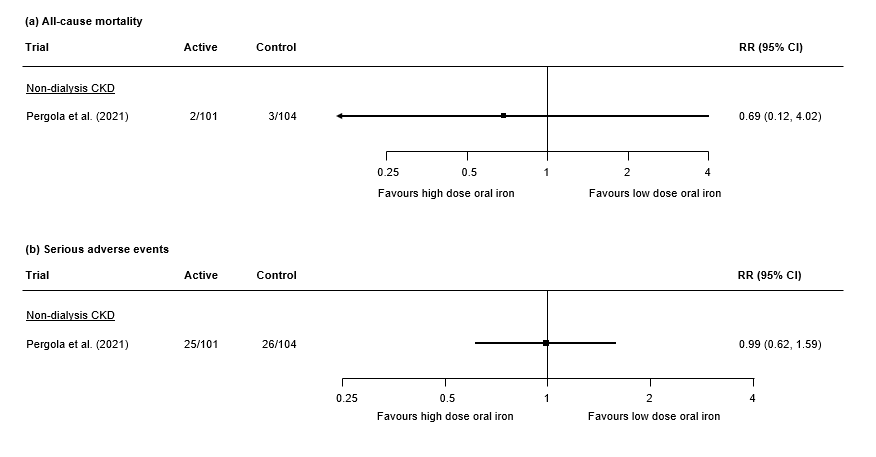

CKD: chronic kidney disease; RR: relative risk; CI: confidence interval.

**Table S1. Characteristics of included trials comparing intravenous vs. oral iron therapies.**

| Trial | Year | No. of Participants with CKD / Total Study Participants | Duration of Follow-up | Primary Population | Intervention | Comparator | Mean Age | Proportion Female (%) | Primary Outcome |
| --- | --- | --- | --- | --- | --- | --- | --- | --- | --- |
| Aggarwal et al.^1^ | 2003 | 40 / 40 | 3 months | Non-dialysis CKD | IV iron dextran | PO ferrous sulfate | NR | 27.5 | Change in Hb, packed cell volume and reticulocyte count |
| VanWyck et al.^2^ | 2005 | 182 / 182 | 56 days | Non-dialysis CKD | IV iron sucrose | PO ferrous sulfate | 63.1 | 67.7 | Proportion of patients with Hb rise ≥1.0 g/dL |
| Agarwal et al.^3^ | 2006 | 75 / 75 | 43 days | Non-dialysis CKD | IV sodium ferric gluconate complex | PO ferrous sulfate | 63.8 | 58.7 | Change in Hb |
| McMahon et al.^4^ | 2010 | 85 / 85 | 12 months | Non-dialysis CKD | IV iron sucrose | PO ferrous sulfate | 69.0 | 27.0 | Change in Hb |
| FIND-CKD^5^ | 2014 | 626 / 626 | 56 weeks | Non-dialysis CKD | IV ferric carboxymaltose | PO ferrous sulfate | 69.1 | 62.2 | Time to initiation of ESA, other iron therapy or blood transfusion or 2 consecutive values Hb<10g/dL |
| REVOKE^6^ | 2015 | 136 / 136 | 24 months | Non-dialysis CKD | IV iron sucrose | PO ferrous sulfate | 65.5 | 23.5 | Slope of measured GFR change |
| Pisani et al.^7^ | 2015 | 99 / 99 | 3 months | Non-dialysis CKD | IV iron gluconate | PO pyrophosphate liposomal iron | 51.3 | 71.7 | Change in Hb |
| Kalra et al.^8^ | 2016 | 351 / 351 | 4 weeks | Non-dialysis CKD | IV ferric derisomaltose | PO ferrous sulfate | 57.7 | 55.6 | Change in Hb |

CKD: Chronic kidney disease; IV: intravenous; PO: per oral; Hb: haemoglobin; ESA: erythropoietin stimulating agent; GFR: glomerular filtration rate.

**Table S2. Characteristics of included trials comparing newer vs. older iron formulations.**

| Trial | Year | No. of Participants with CKD | Duration of Follow-up | Primary Population | Intervention | Comparator | Mean Age | Proportion Female (%) | Primary Outcome |
| --- | --- | --- | --- | --- | --- | --- | --- | --- | --- |
| Kosch et al.^9^ | 2001 | 59 / 59 | 6 months | Hemodialysis | IV iron sucrose | IV iron gluconate | NR | NR | Change in Hb |
| Agarwal et al.^10^ | 2011 | 62 / 62 | 36 days | Non-dialysis CKD | IV ferric gluconate | IV iron sucrose | 65.3 | 58.1 | Urine protein to creatinine ratio |
| HEMATOCRIT^11^ | 2012 | 62 / 62 | 6 months | Peritoneal dialysis | PO heme iron polypeptide | PO ferrous sulfate | 59.5 | 57.4 | Transferrin saturation |
| REPAIR-IDA^12^ | 2014 | 2584 / 2584 | 56 days | Non-dialysis CKD | IV ferric carboxymaltose | IV iron sucrose | 67.3 | 63.6 | Change in Hb |
| Bhandari et al.^13^ | 2015 | 351 / 351 | 6 weeks | Hemodialysis | IV ferric derisomaltose | IV iron sucrose | 59.9 | 34.2 | Proportion of patients with Hb 9.5-12.5 g/dL at 6 weeks |
| FACT^14^ | 2019 | 293 / 293 | 11 months | Hemodialysis | IV ferumoxytol | IV iron sucrose | 58.7 | 41.6 | Change in Hb |
| COPEFER^15^ | 2021 | 142 / 142 | 40 weeks | Hemodialysis | IV ferric carboxymaltose | IV iron sucrose | 58.5 | 28.9 | Change in Hb |
| IRON-CKD^16^ | 2021 | 40 / 40 | 3 months | Non-dialysis CKD | IV ferric derisomaltose | IV iron sucrose and iron dextran | 58.8 | 42.5 | Oxidative stress (measured by thiobarbituric acid reactive substances and labile plasma iron) and inflammation (measured by interleukin and CRP levels) |
| FERWON-NEPHRO^17^ | 2021 | 1538 / 1538 | 8 weeks | Non-dialysis CKD | IV ferric derisomaltose | IV iron sucrose | 68.6 | 62.5 | Change in Hb and serious or severe hypersensitivity reactions |

CKD: Chronic kidney disease; IV: intravenous; PO: per oral; Hb: haemoglobin; CRP: C-reactive protein.

**Table S3. Characteristics of included trials comparing higher vs. lower dose oral iron therapies**

| Trial | Year | No. of Participants with CKD | Duration of Follow-up | Primary Population | Intervention | Comparator | Mean Age | Proportion Female (%) | Primary Outcome |
| --- | --- | --- | --- | --- | --- | --- | --- | --- | --- |
| Pergola et al.^18^ | 2021 | 206 / 206 | 48 weeks | Non-dialysis CKD | PO ferric citrate | PO ferric citrate | 69.5 | 64.3 | Change in Hb |
| Sood et al.^19^ | 2023 | 80 / 80 | 12 weeks | Non-dialysis CKD | PO ferric citrate | PO ferric citrate | 50.0 | 30.0 | Change in transferrin saturation |

CKD: Chronic kidney disease; IV: intravenous; PO: per oral; Hb: haemoglobin.

**Table S4. Risk of bias assessment – iron vs. usual care or placebo.**

| **Trial** | **Randomisation process** | **Deviations from intended interventions** | **Missing outcome data** | **Outcome measurement** | **Selection of reported result** | **Overall** |
| --- | --- | --- | --- | --- | --- | --- |
| Besarab et al.^20^ | Some concerns | Low | Low | Low | Low | Some concerns |
| DRIVE^21^ | Low | Low | Low | Some concerns | Some concerns | Some concerns |
| FAIR-HF^22^ | Low | Low | Low | Low | Low | Low |
| McIntyre et al.^23^ | Low | Low | Low | Some concerns | Low | Some concerns |
| Charytan et al.^24^ | Low | Low | Low | Low | Low | Low |
| Yokoyama et al.^25^ | Low | Low | Low | Low | Low | Low |
| Block et al.^26^ | Low | Low | Low | Low | Low | Low |
| CONFIRM-HF^27^ | Low | Low | Low | Low | Low | Low |
| CRUISE 1 and 2^27^ | Low | Some concerns | Low | Low | Low | Some concerns |
| PRIME^28^ | Low | Low | Low | Low | Low | Low |
| Floege et al.^29^ | Low | Low | Low | Low | Low | Low |
| Lewis et al.^30^ | Low | Some concerns | Low | Low | Low | Some concerns |
| Fishbane et al.^31^ | Low | Low | Low | Low | Low | Low |
| Koiwa et al.^32^ | Low | Low | Low | Some concerns | Low | Some concerns |
| Iguchi et al.^33^ | High | High | High | Low | Some concerns | High |
| Block et al.^34^ | Low | Some concerns | Low | Some concerns | Low | Some concerns |
| PIVOTAL^35^ | Low | Low | Low | Low | Low | Low |
| AFFIRM-AHF^36^ | Low | Low | Low | Low | Low | Low |
| van den Oever et al.^37^ | Low | High | Some concerns | Low | Low | High |
| Susantitaphong et al.^38^ | Low | Some concerns | Low | Low | Low | Some concerns |
| AEGIS-CKD^39^ | Low | Low | Low | Low | Low | Low |
| ZununiVahed et al.^40^ | Some concerns | Low | Low | Low | Low | Some concerns |
| The Iron and the Heart Study^41^ | Some concerns | Low | Low | Low | Low | Some concerns |
| IRONMAN^42^ | Low | Low | Low | Low | Low | Low |
| HEART-FID^43^ | Low | Low | Low | Low | Low | Low |
| MAINTAIN-IRON^44^ | Low | Low | Low | Low | Low | Low |

**Table S5. Risk of bias assessment – intravenous vs. oral iron.**

| **Trial** | **Randomisation process** | **Deviations from intended interventions** | **Missing outcome data** | **Outcome measurement** | **Selection of reported result** | **Overall** |
| --- | --- | --- | --- | --- | --- | --- |
| Aggarwal et al.^1^ | Some concerns | High | Low | Some concerns | Low | High |
| VanWyck et al.^2^ | Some concerns | Some concerns | Low | Some concerns | Low | Some concerns |
| Agarwal et al.^3^ | Low | Low | Low | Some concerns | High | High |
| McMahon et al.^4^ | Low | High | Some concerns | Low | Low | High |
| FIND-CKD^5^ | Low | Low | Low | Low | Low | Low |
| REVOKE^6^ | Low | High | Low | Some concerns | Low | High |
| Pisani et al.^7^ | Low | Some concerns | Low | High | Low | High |
| Kalra et al.^8^ | Low | Some concerns | Low | High | Low | High |

**Table S6. Risk of bias assessment – newer generation iron vs. older generation iron formulations.**

| **Trial** | **Randomisation process** | **Deviations from intended interventions** | **Missing outcome data** | **Outcome measurement** | **Selection of reported result** | **Overall** |
| --- | --- | --- | --- | --- | --- | --- |
| Kosch et al.^9^ | Some concerns | Some concerns | Low | High | Low | High |
| Agarwal et al.^10^ | Low | Low | Low | Low | High | High |
| HEMATOCRIT^11^ | Low | Low | Low | Some concerns | Low | Some concerns |
| REPAIR-IDA^12^ | Low | Low | Low | Low | Low | Low |
| Bhandari et al.^13^ | Low | Low | Low | Some concerns | Low | Some concerns |
| FACT^14^ | Low | Low | Low | Some concerns | Low | Some concerns |
| COPEFER^15^ | Low | Low | Some concerns | Low | Low | Some concerns |
| IRON-CKD^16^ | Low | High | Low | Some concerns | Low | High |
| FERWON-NEPHRO^17^ | Low | Low | Low | Low | Low | Low |

**Table S8. Changes in eGFR (mL/min/1.73m^2^) with iron therapy vs. usual care or placebo.**

| Trial | Iron | | Usual care or placebo | |
| --- | --- | --- | --- | --- |
|  | Mean (SD), baseline | Mean (SD), follow-up | Mean (SD), baseline | Mean (SD), follow-up |
| The Iron and the Heart Study^41^ | 33.2 (9.3) | 32.1 (9.5) | 29.1 (9.9) | 28.2 (8.6) |
| Iguchi et al.^33^ | 24.4 (9.8) | 25.0 (11.2) | 31.3 (10.1) | 32.1 (11.0) |
| Yokoyama et al.^25^ | 8.6 (3.9) | 7.9 (4.3) | 9.8 (8.2) | 9.0 (7.3) |
| FAIR-HF^22^ | 7.1 ± 1.24 (SE) | | 5.56 ± 1.74 (SE) | |

SD: standard deviation; SE: standard error.

**Table S9. Changes in eGFR (mL/min/1.73m^2^) with intravenous vs. oral iron.**

| Trial | Intravenous iron | | Oral iron | |
| --- | --- | --- | --- | --- |
|  | Mean (SD), baseline | Mean (SD), follow-up | Mean (SD), baseline | Mean (SD), follow-up |
| REVOKE^6^ | 34.3 (10.2) | NR | 34.7 (10) | NR |
|  | -4 per year (NR) | | -3.6 per year (NR) | |
|  | Between group differences: -0.35 (95% CI -2.9 to 2.3) | | | |
| Pisani et al.^7^ | 31.8 (12.9) | 27.9 (7.8) | 25.9 (11.4) | 25.1 (12.7) |
| FIND-CKD^5^ | 32.0 (1.1) | 33.2 (1.4) | 33.2 (0.8) | 33.7 (1.0) |
|  | -0.6 (0.8) | | -1.1 (0.6) | |
| McMahon et al.^4^ | 25.0 (8.0) | 23.0 (8.0) | 26.0 (11.0) | 22.0 (10.0) |
| VanWyck et al.^2^ | 30.4 (NR) | NR | 28.5 (NR) | NR |
|  | -1.45 (95% CI -2.67 to -0.2) | | -4.4 (95% CI -6.29 to -2.5) | |

SD: standard deviation; CI: confidence interval. NR: not reported.

**Table S10. Changes in eGFR (mL/min/1.73m^2^) with higher vs. lower dose oral iron.**

| Trial | Higher dose oral iron | | Lower dose oral iron | |
| --- | --- | --- | --- | --- |
|  | Mean (SD), baseline | Mean (SD), follow-up | Mean (SD), baseline | Mean (SD), follow-up |
| Sood et al.^19^ | 33.8 (11.1) | 34.3 (16.3) | 39.5 (12.4) | 38.6 (15.7) |
| Pergola et al.^18^ | 34.5 (11.6) | NR | 32.8 (10.1) | NR |
|  | -2.20 (95% CI -4.24 to -0.15) | | -1.85 (95% CI -3.78 to -0.08) | |

SD: standard deviation; CI: confidence interval; NR: not reported.

**Table S11. Changes in proteinuria (mg/g) with iron therapy vs. usual care or placebo.**

| Trial | Iron | | Usual care or placebo | |
| --- | --- | --- | --- | --- |
|  | Mean (SD), baseline  (mg/g) | Mean (SD), follow-up (mg/g) | Mean (SD), baseline  (mg/g) | Mean (SD), follow-up (mg/g) |
| The Iron and the Heart Study^41^ | 458.8 (524.2) | 513.6 (815.9) | 996.3 (1456.8) | 618.8 (902.6) |

SD: standard deviation.

**Table S12. Changes in proteinuria with newer vs older generation iron.**

| Trial | On ACEI / ARB | | No ACEI/ARB | |
| --- | --- | --- | --- | --- |
| Agarwal et al.^10^ | Newer generation iron (% change in UPCR from baseline) | Older generation iron (% change in UPCR from baseline) | Newer generation iron (% change in UPCR from baseline) | Older generation iron (% change in UPCR from baseline) |
|  | NR | NR | NR | NR |
|  | Iron sucrose (older generation) produced 78% higher UPCR compared to ferric gluconate (newer generation) | | Iron sucrose (older generation) produced 40.9% higher UPCR compared to ferric gluconate (newer generation) | |

NR: not reported.

**Table S13. Changes in proteinuria (mg/g) with higher vs. lower dose oral iron.**

| Trial | Higher dose | | Lower dose | |
| --- | --- | --- | --- | --- |
|  | Median (IQR), baseline (mg/g) | Median (IQR), follow-up (mg/g) | Median (IQR), baseline (mg/g) | Median (IQR), follow-up (mg/g) |
| Sood et al.^19^ | 0.54 (0.17-1.74) | 0.37 (0.24-1.13) | 0.30 (0.13-0.95) | 0.46 (0.205-1.00) |

IQR: interquartile range.

**Table S14. Changes in albuminuria (mg/g) with iron therapy vs. usual care or placebo.**

| Trial | Iron | | Usual care or placebo | |
| --- | --- | --- | --- | --- |
|  | Mean (SD), baseline  (mg/g) | Mean (SD), follow-up (mg/g) | Mean (SD), baseline  (mg/g) | Mean (SD), follow-up (mg/g) |
| The Iron and the Heart Study^41^ | 237.8 (353.6) | 437.6 (633.8) | 838.0 (1603.6) | 375.7 (395.1) |

**Table S15. Changes in albuminuria with newer vs older generation iron.**

| Trial | On ACEI / ARB | | No ACEI/ARB | |
| --- | --- | --- | --- | --- |
| Agarwal et al.^10^ | Newer generation iron (% change in UACR from baseline) | Older generation iron (% change in UACR from baseline) | Newer generation iron (% change in UACR from baseline) | Older generation iron (% change in UACR from baseline) |
|  | NR | NR | NR | NR |
|  | Iron sucrose (older generation) produced 135% higher UACR compared to ferric gluconate (newer generation) | | NR | |

NR: not reported.

| **Section and Topic** | **Item #** | **Checklist item** | **Location where item is reported** |
| --- | --- | --- | --- |
| **TITLE** | | |  |
| Title | 1 | Identify the report as a systematic review. | Page 1 |
| **ABSTRACT** | | |  |
| Abstract | 2 | See the PRISMA 2020 for Abstracts checklist. | Page 2 |
| **INTRODUCTION** | | |  |
| Rationale | 3 | Describe the rationale for the review in the context of existing knowledge. | Page 4 |
| Objectives | 4 | Provide an explicit statement of the objective(s) or question(s) the review addresses. | Page 5 |
| **METHODS** | | |  |
| Eligibility criteria | 5 | Specify the inclusion and exclusion criteria for the review and how studies were grouped for the syntheses. | Page 6 |
| Information sources | 6 | Specify all databases, registers, websites, organisations, reference lists and other sources searched or consulted to identify studies. Specify the date when each source was last searched or consulted. | Page 5 |
| Search strategy | 7 | Present the full search strategies for all databases, registers and websites, including any filters and limits used. | Table S1 |
| Selection process | 8 | Specify the methods used to decide whether a study met the inclusion criteria of the review, including how many reviewers screened each record and each report retrieved, whether they worked independently, and if applicable, details of automation tools used in the process. | Page 6 |
| Data collection process | 9 | Specify the methods used to collect data from reports, including how many reviewers collected data from each report, whether they worked independently, any processes for obtaining or confirming data from study investigators, and if applicable, details of automation tools used in the process. | Page 6-7 |
| Data items | 10a | List and define all outcomes for which data were sought. Specify whether all results that were compatible with each outcome domain in each study were sought (e.g. for all measures, time points, analyses), and if not, the methods used to decide which results to collect. | Page 7 |
|  | 10b | List and define all other variables for which data were sought (e.g. participant and intervention characteristics, funding sources). Describe any assumptions made about any missing or unclear information. | Page 6-8 |
| Study risk of bias assessment | 11 | Specify the methods used to assess risk of bias in the included studies, including details of the tool(s) used, how many reviewers assessed each study and whether they worked independently, and if applicable, details of automation tools used in the process. | Page 6-7 |
| Effect measures | 12 | Specify for each outcome the effect measure(s) (e.g. risk ratio, mean difference) used in the synthesis or presentation of results. | Page 7 |
| Synthesis methods | 13a | Describe the processes used to decide which studies were eligible for each synthesis (e.g. tabulating the study intervention characteristics and comparing against the planned groups for each synthesis (item #5)). | Page 7-8 |
|  | 13b | Describe any methods required to prepare the data for presentation or synthesis, such as handling of missing summary statistics, or data conversions. | Page 7-8 |
|  | 13c | Describe any methods used to tabulate or visually display results of individual studies and syntheses. | Page 8 |
|  | 13d | Describe any methods used to synthesize results and provide a rationale for the choice(s). If meta-analysis was performed, describe the model(s), method(s) to identify the presence and extent of statistical heterogeneity, and software package(s) used. | Page 8 |
|  | 13e | Describe any methods used to explore possible causes of heterogeneity among study results (e.g. subgroup analysis, meta-regression). | Page 8 |
|  | 13f | Describe any sensitivity analyses conducted to assess robustness of the synthesized results. | Page 8 |
| Reporting bias assessment | 14 | Describe any methods used to assess risk of bias due to missing results in a synthesis (arising from reporting biases). | Page 7 |
| Certainty assessment | 15 | Describe any methods used to assess certainty (or confidence) in the body of evidence for an outcome. | Page 8 |
| **RESULTS** | | |  |
| Study selection | 16a | Describe the results of the search and selection process, from the number of records identified in the search to the number of studies included in the review, ideally using a flow diagram. | Page 8 |
|  | 16b | Cite studies that might appear to meet the inclusion criteria, but which were excluded, and explain why they were excluded. | N/A |
| Study characteristics | 17 | Cite each included study and present its characteristics. | Table 1, S1-3 |
| Risk of bias in studies | 18 | Present assessments of risk of bias for each included study. | Table S4-7 |
| Results of individual studies | 19 | For all outcomes, present, for each study: (a) summary statistics for each group (where appropriate) and (b) an effect estimate and its precision (e.g. confidence/credible interval), ideally using structured tables or plots. | Figures 1-5, S2-4 |
| Results of syntheses | 20a | For each synthesis, briefly summarise the characteristics and risk of bias among contributing studies. | Page 9 |
|  | 20b | Present results of all statistical syntheses conducted. If meta-analysis was done, present for each the summary estimate and its precision (e.g. confidence/credible interval) and measures of statistical heterogeneity. If comparing groups, describe the direction of the effect. | Page 9-10, Figures 1-5, S2-4, |
|  | 20c | Present results of all investigations of possible causes of heterogeneity among study results. | Page 9-10 |
|  | 20d | Present results of all sensitivity analyses conducted to assess the robustness of the synthesized results. | N/A |
| Reporting biases | 21 | Present assessments of risk of bias due to missing results (arising from reporting biases) for each synthesis assessed. | N/A |
| Certainty of evidence | 22 | Present assessments of certainty (or confidence) in the body of evidence for each outcome assessed. | Page 9-10, Figures 1-5, S2-4 |
| **DISCUSSION** | | |  |
| Discussion | 23a | Provide a general interpretation of the results in the context of other evidence. | Page 10-11 |
|  | 23b | Discuss any limitations of the evidence included in the review. | Page 13-14 |
|  | 23c | Discuss any limitations of the review processes used. | Page 13-14 |
|  | 23d | Discuss implications of the results for practice, policy, and future research. | Page 13-15 |
| **OTHER INFORMATION** | | |  |
| Registration and protocol | 24a | Provide registration information for the review, including register name and registration number, or state that the review was not registered. | Page 5 |
|  | 24b | Indicate where the review protocol can be accessed, or state that a protocol was not prepared. | Page 5 |
|  | 24c | Describe and explain any amendments to information provided at registration or in the protocol. | N/A |
| Support | 25 | Describe sources of financial or non-financial support for the review, and the role of the funders or sponsors in the review. | Page 15 |
| Competing interests | 26 | Declare any competing interests of review authors. | Page 15 |
| Availability of data, code and other materials | 27 | Report which of the following are publicly available and where they can be found: template data collection forms; data extracted from included studies; data used for all analyses; analytic code; any other materials used in the review. | Page 1 |

*From:* Page MJ, McKenzie JE, Bossuyt PM, Boutron I, Hoffmann TC, Mulrow CD, et al. The PRISMA 2020 statement: an updated guideline for reporting systematic reviews. BMJ 2021;372:n71. doi: 10.1136/bmj.n71

For more information, visit: <http://www.prisma-statement.org/>
